# Feature Selection for an Explainability Analysis in Detection of COVID-19 Active Cases from Facebook User-Based Online Surveys

**DOI:** 10.1101/2023.05.26.23290608

**Authors:** Jesús Rufino, Juan Marcos Ramírez, Jose Aguilar, Carlos Baquero, Jaya Champati, Davide Frey, Rosa Elvira Lillo, Antonio Fernández-Anta

## Abstract

In this paper, we introduce a machine-learning approach to detecting COVID-19-positive cases from self-reported information. Specifically, the proposed method builds a tree-based binary classification model that includes a recursive feature elimination step. Based on Shapley values, the recursive feature elimination method preserves the most relevant features without compromising the detection performance. In contrast to previous approaches that use a limited set of selected features, the machine learning approach constructs a detection engine that considers the full set of features reported by respondents. Various versions of the proposed approach were implemented using three different binary classifiers: random forest (RF), light gradient boosting (LGB), and extreme gradient boosting (XGB). We consistently evaluate the performance of the implemented versions of the proposed detection approach on data extracted from the University of Maryland Global COVID-19 Trends and Impact Survey (UMD-CTIS) for four different countries: Brazil, Canada, Japan, and South Africa, and two periods: 2020 and 2021. We also compare the performance of the proposed approach to those obtained by state-of-the-art methods under various quality metrics: F1-score, sensitivity, specificity, precision, receiver operating characteristic (ROC), and area under ROC curve (AUC). It should be noted that the proposed machine learning approach outperformed state-of-the-art detection techniques in terms of the F1-score metric. In addition, this work shows the normalized daily case curves obtained by the proposed approach for the four countries. It should note that the estimated curves are compared to those reported in official reports. Finally, we perform an explainability analysis, using Shapley and relevance ranking of the classification models, to identify the most significant variables contributing to detecting COVID-19-positive cases. This analysis allowed us to determine the relevance of each feature and the corresponding contribution to the detection task.

## 1. Introduction

During the global pandemic of Coronavirus disease 2019 (COVID-19) caused by the spread of the severe acute respiratory syndrome Coronavirus 2 (SARS-CoV-2) [1], healthcare systems faced the challenge of devising surveillance strategies to monitor the disease expansion. Particularly, these strategies required high-quality data in almost real-time to design the healthcare policies [2]. In this sense, massive screening using the polymerase chain reaction (PCR) test has been the most widely used approach for monitoring disease expansion. Nevertheless, the accuracy of COVID-19 detection from PCR tests is impacted by several factors, including the timing of the test relative to the infection [3], the high rate of asymptomatic cases [4], and the limited availability of test kits [5]. Therefore, various approaches that leverage survey data have been used to provide supplementary information for tracking pandemic indicators. For instance, symptoms provided by individuals tested via PCR have been analyzed to evaluate COVID-19 detection methods [6, 5]. Similar approaches based on smartphone apps have also been developed to detect infected individuals from the self-report symptoms [7, 8, 9]. Online questionnaires about symptoms, social behavior, and isolation measures have also been publicized through social networks [10] to detect positive cases.

Several methods have been proposed for COVID-19 detection based on individual features extracted from surveys. These detection methods can be categorized into three groups: prediction rules, logistic regression methods, and machine learning models. Prediction rule methods declare an active case based on a specific set of reported symptoms. COVID-like illness (CLI) approved by either the Centers for Disease Control and Prevention (CDC) or the World Health Organization (WHO) are the most representative prediction rules [11, 12, 13]. Additional prediction rules have been presented in [14, 15, 16]. Logistic regression techniques build a linear expression whose parameters represent the contribution of the reported features (symptom, gender, age group) [6, 7, 10, 17, 18, 19]. These techniques typically use a reduced set of (usually less than five) features to identify active cases [2, 5]. These methods ignore information provided by features other than symptoms and have low detection accuracy.

In April 2020, the University of Maryland (UMD), in collaboration with Facebook, launched the Global COVID-19 Trends and Impact Survey (UMD-CTIS), a large health surveillance system based on surveys [20, 2]. More precisely, the purpose of this study was to gather daily information from a representative sample of Facebook’s Active User Base (FAUB), who were invited to participate in the survey to consult them about various COVID-19-related characteristics, including symptoms, PCR test outcomes, vaccination acceptance, isolation measures, and demographics. Questionnaires were translated into 56 languages, and data were collected from 114 countries/territories, reaching a wide range of social and economic groups. Note also that the UMD-CTIS data provide fine-grained coverage of pandemic trends, which permits estimating various health indicator trends for different regions.

In this paper, we introduce a machine-learning approach that includes tree-based supervised classifiers and feature-selection strategies to detect COVID-19-active cases. Contrary to previous methods, the proposed approach considers a broad range of individual characteristics. To reduce the dimensionality of the input data and minimize overfitting risk, the training stage of the supervised classifiers includes a feature-selection technique founded on Shapley values that determines the optimal set of features to achieve the best balance between model complexity and detection accuracy [21]. Six versions of the proposed approach were implemented with three different tree-based classifiers: random forest (RF), light gradient boosting (LGB), and extreme gradient boosting (XGB). This approach is evaluated using UMD-CTIS data from four countries: Brazil, Canada, Japan, and South Africa, and for 2020 and 2021. The performance of the proposed approach is evaluated using multiple quality metrics. In addition, we compare the performance of the developed machine learning approach against those produced by other state-of-the-art techniques. In general, our approach outperforms other methods in terms of the F1-score. We also compare COVID-19 daily estimates obtained by machine-learning implementations with those yielded by official reports. Furthermore, this work performs an explainability analysis that identifies the relevant input features and describes how they contribute to the detection task[22, 23]. The paper is organized as follows. Section 2 describes the materials and methods, and the results are displayed in Section 3. The discussion generated by the explainability analysis is in Section 4.

## 2. Materials and methods

In this section, we describe the UMD-CTIS dataset, machine learning methods used to identify COVID-19-positive cases, and the feature selection technique used.

### 2.1. Study population

We evaluate the performance of the proposed detection approach using datasets extracted from the UMD-CTIS collected for four countries: Brazil (BR), Canada (CA), Japan (JP), and South Africa (ZA). Geographic diversity and the availability of sufficient data samples were considered when selecting these countries. In addition, we evaluate the detection methods for two periods: 2020 (from April 23 to December 31, 2020), and 2021 (from January 1 to December 31, 2021). We selected these periods to observe the impact of the vaccination campaigns on the selection of relevant features during the training stage and the estimation accuracy. Afterward, for each country and period, we extract responses from participants who have reported at least one symptom during the previous 24 hours and have provided a test result within the past 14 days. In contrast to previous approaches that take into account a reduced set of individual features, our approach considers the full set of features collected by the UMD-CTIS questionnaires.

A summary of the study population characteristics is provided in Table 1 for the four countries and 2020 and 2021. For 2020, we analyze 104,746 respondents (BR: 83,238, CA: 8,927, JP: 4,698, and ZA: 7,883) who reported at least one symptom in the last 24 hours and provided a test result within the past 14 days (Tested symptomatic). Similarly, we extracted data from 370,728 individuals in 2021 (BR: 262,683, CA: 33,997, JP: 41,010, and ZA: 23,038 ZA). Table 1 also includes the number of *positive* and *negative tests* among *tested symptomatic*, as well as the *test positive rate* (TPR), calculated as follows: TPR = (100×positive)∕(Tested symptomatic), for each country and period. Note that the TPR values obtained for Brazil and South Africa are at least three times larger than those yielded by Canada and Japan for 2020 and 2021. Table 1 displays information on other individual features such as gender, age group, the average number of reported symptoms per questionnaire, and the average number of reported symptoms per questionnaire among positives. Figure 1 depicts the percentage of tested positives who reported a particular symptom for each country and period in descending order. In addition, Figure 1 illustrates the corresponding rate of tested symptomatic reporting each symptom. As can be seen in this figure, fatigue is the most common symptom among positives, with the highest rates in the bar plots. In general, the symptom patterns vary among the various countries and for 2020 and 2021.

**Figure 1:**
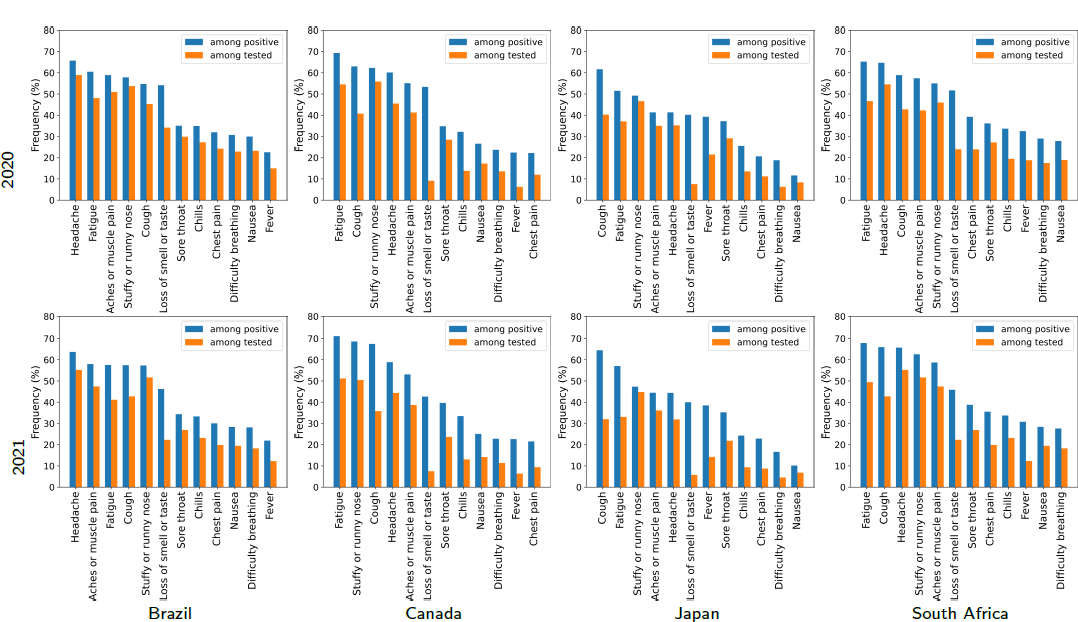
Rate of positive who reported a particular symptom for the four countries and 2020 and 2021. The bar plots also include the rate of test symptomatic reporting each symptom.

**Table 1.** Characteristics of the study population for the various countries and two non-overlapped periods (2020 and 2021).

| Characteristic | Brazil |  | Canada |  | Japan |  | South Africa |  |
| --- | --- | --- | --- | --- | --- | --- | --- | --- |
|  | 2020 | 2021 | 2020 | 2021 | 2020 | 2021 | 2020 | 2021 |
| 1. Tested symptomatic | 83238 | 262683 | 8927 | 33997 | 4698 | 41010 | 7883 | 23038 |
| 2. Test outcome |  |  |  |  |  |  |  |  |
| (a) Positive | 44963 | 106471 | 838 | 3433 | 532 | 4011 | 2866 | 8459 |
| (b) Negative | 38275 | 156212 | 8089 | 30564 | 4166 | 36999 | 5017 | 14579 |
| (c) TPR | 54.02% | 40.53% | 9.39% | 10.10% | 11.32% | 9.78% | 36.35% | 36.71% |
| 3. Gender |  |  |  |  |  |  |  |  |
| (a) Female | 45357 | 130235 | 5438 | 19472 | 1679 | 14283 | 3923 | 11291 |
| (b) Male | 24928 | 76689 | 2315 | 9824 | 2388 | 20791 | 2525 | 6730 |
| 4. Age groups |  |  |  |  |  |  |  |  |
| (a) 18-24 | 8270 | 27474 | 1136 | 3248 | 179 | 871 | 739 | 1580 |
| (b) 25-34 | 19596 | 56227 | 2337 | 7172 | 577 | 3797 | 2252 | 4889 |
| (c) 35-44 | 21061 | 57452 | 1750 | 6688 | 997 | 7527 | 1801 | 4721 |
| (d) 45-54 | 13776 | 39122 | 1210 | 5215 | 1216 | 10413 | 1141 | 3878 |
| (e) 55-64 | 6968 | 22190 | 954 | 4478 | 828 | 8724 | 491 | 2124 |
| (f) 65-74 | 140 | 6016 | 308 | 2421 | 479 | 3529 | 1667 | 799 |
| (g) 75+ | 233 | 1025 | 126 | 825 | 66 | 846 | 27 | 230 |
| 5. Average number of reported symptoms | 4.33 | 3.80 | 3.38 | 3.07 | 2.92 | 2.49 | 3.82 | 3.95 |
| 6. Average number of reported symptoms among positive | 5.37 | 5.16 | 5.25 | 5.27 | 4.38 | 4.45 | 5.51 | 5.61 |

### 2.2. Machine Learning Methods

Machine-learning (ML) models used in this work are tree-based classifiers [24, 25]. Specifically, we will use random forest (RF), extreme gradient boosting (XGB), and light gradient boosting (LGB). These models provide relevance rankings of the input variables, which serves as the basis for performing an explainability analysis process. Now, each model is presented:

- **Random Forest (RF)** [26, 24, 25]: RF consists of a set of decision trees, each generated by a bagging algorithm. These trees form a “forest” of trees voting for a specific result. This algorithm uses bootstrapping to fit decision trees with sub-samples of the original dataset. For each tree, the algorithm uses averaging to improve predictive accuracy and control overfitting. In classification algorithms, the most common output will be chosen as the final output of the algorithm.
- **Extreme Gradient-Boosting (XGB)** [27, 24, 25]: It is a class of ensemble machine-learning algorithms that can be used for classification problems. Ensembles are constructed from decision-tree models. The trees are added one at a time to the ensemble, and fitted to correct for prediction errors made by previous models. This is done using data that could not be learned so far. This technique is known as boosting. Moreover, XGB applies a regularization technique to reduce overfitting.
- **Light Gradient-Boosting (LGB)** [27, 24, 25]: LGB is a gradient-boosting algorithm based on decision trees which decreases memory usage and improves model efficiency. It uses two techniques: the classic XGB based on Exclusive Feature Bundling, and the Gradient-driven One Side Sampling (GOSS). GOSS keeps those instances with large gradients (they will contribute more to information gain) and randomly drops those instances with small gradients to improve accuracy estimation. It is faster than the XGB algorithm because GOSS filters out the data instances to find a suitable split value. This makes the process faster.

### 2.3. Explainability Analysis Methods

In medicine, there is an increasing demand for AI approaches that are both efficient and transparent, as well as easily explainable by a human expert[22, 23]. Currently, it is difficult to find explanations as to why a result occurs or how a model describes the underlying biological process [28]. In COVID-19 studies that use machine learning models, explainable AI is urgently needed to understand and retrace the machine’s decision-making process. It is critical, for example, to analyze the relationships between symptoms, age groups, gender, and COVID-19 cases. On the other hand, explaining, interpreting, or understanding, are synonymous in the context of explainable AI, and various approaches have been proposed [28]. As a first categorization, the explanator must describe the model or result (e.g., classification, prediction). Therefore, this classification is whether the explicability is *global* to provide insights into the inner workings of the entire model for some specific dataset or *local* for a single test input *x* and its corresponding result *y* [29, 28]. There are two types of explicability: *ante-hoc* consists of building it directly from the beginning of model creation (the model can be understood immediately), and *post-hoc* consists of building it after model creation using a technique to extract the explanation [29, 28].

Global explanators try to reveal certain properties of the model independently of results. An example is the tree-based approach (e.g., decision trees and ensembles of decision trees, such as RF) [26, 30, 27]. In this case, the information gained from a variable accumulated over all trees can be used as a relevance measure. Another example of a feature importance metric for tree-based methods is the feature’s depth in the tree. Local explanations are only valid near a result. The classic methods relate the model result to the feature vector by ranking the explanatory power, i.e. the salience of each feature. There are two main families of methods. The first, *Attention Based Models*, examines the most promising parts of input features that lead to a certain output for a given task. For a given output, they try to find out whether input features with high attention weights were responsible for the outcome. The second is *feature-attribution approaches* that explicitly try to quantify the contribution of each individual feature to the results. In this work, we analyze a global ante-hoc approach based on the tree-based methods using the feature ranking provided by them and a local and post-hoc approach that extracts the feature importance for a given input based on the fact that output can be written as the sum of bias and single feature contributions (Shapley values).

- **SHapley Additive exPlanations (SHAP values) [21, 31]:** Shapley values are an example of local approaches [21, 31], which are created by means of a method from coalitional game theory, assuming that each feature value of the instance is a player in a game where the output is the payoff. Shapley values dispense the payoff among the features. The goal of SHAP is to explain the output of a model by computing the contribution of each feature to this output. For that, SHAP computes Shapley values. A Shapley value assigns each feature an importance value for a particular output to define the explanation. This value, for feature *i*, is the unified measure of additive feature attributions (*ϕ*_*i*_) [21, 31]:

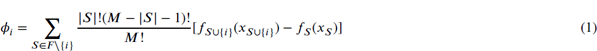

Where ***F*** is the set of input features, ***S*** is a subset of input features, ***M*** is the number of input features and *f*_***S***_ (.) represents the output function of the model (e.g. prediction). In this equation, 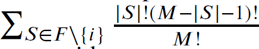 represents the weighted average of all possible subsets of ***S*** in ***F***. In addition, this equation considers the difference between when this feature is present in the output (*f*_***S***∪{*i*}_(*x*_***S***∪{*i*}_) and when it is absent (*f*_***S***_(*x*_***S***_)). With these SHAP values, we are able to select the variables that give the model the highest contribution. The calculation time increases exponentially according to the number of features. To avoid this, one solution is to determine contributions for only a few samples of the possible coalitions. We have used a sample of 100 to compute the importance of each feature using the kernel-explainer function.

- **Tree-based methods [26, 30]:** The explainability analysis for tree-based ML methods is possible by their capabilities to rank their features/variables. These methods make it possible to compute various feature/variable importance measures to be used in an explainability analysis. For example, MDA (mean decrease in accuracy) determines feature importance (ranking) as the mean decrease of accuracy over all predictions, when a given variable is permuted after training [26, 30]. Thus, MDA calculates the average decrease of accuracy against random permutations of feature values in different cases. The cases use a trained model, and each tree of the model is permuted along the *m*-th feature and the average of these differences for all decision trees gives the m-th feature’s MDA. Also, it can use the Gini value, which measures the average gain of purity by splits of a given variable [26, 30]. If the variable is useful, then it tends to split labeled nodes into single classes. The permuted variables tend neither to increase nor to decrease node purities. Permuting a useful variable tends to yield a large decrease in mean Gini gain. Gini importance is normally inferior to variable importance because it is more unstable and biased.

Finally, in some cases, the large number of features could be a problem because of the noise produced by some of the variables or their low significance. There are some ML algorithms that already have some regularization mechanisms that reduce the number of features. However, the techniques being used in this study do not have any of these mechanisms. For this reason, we also use SHAP values as a feature-reduction technique that we apply to each of them. Ultimately, we end up with 6 different models, 3 of them with all the features (RF, LGBM and XGB), and 3 with the features selected according to the Shapley values (RF.SHAP, LGBM.SHAP and XGB.SHAP).

## 3. Results

### 3.1. Performance analysis

Table 2 illustrates different performance metrics in percentage and the 95% confidence intervals (CIs) generated by the various implementations of the proposed approach for Brazil, in 2020 and 2021. In particular, this table includes F_1_-score, specificity, sensitivity, and precision obtained by the proposed techniques. A bold font and an underlined value indicate the best and second-best values for each metric and year. For Brazil 2020, the method based on the RF classification method generates the best performance results, i.e., F_1_-score (RF: 84.24%, 95% CI: 84.19% to 84.30%), specificity (RF: 82.88%, 95% CI: 82.79% to 82.98%), sensitivity (RF: 83.43%, 95% CI: 83.35% to 83.51%), and precision (RF: 85.08%, 95% CI: 85.00% to 85.15%). In addition, the second best values are generated by the RF model optimized using the recursive feature elimination stage based on Shapley values: F_1_-score (RF SHAP: 84.23%, 95% CI: 84.18% to 84.28%), specificity (RF SHAP: 82.87%, 95% CI: 82.78% to 82.97%), sensitivity (RF SHAP: 83.41%, 95% CI: 83.33% to 83.49%), and precision (RF SHAP: 85.07%, 95% CI: 84.99% to 85.15%). For 2021, equally, the RF and RF SHAP classifiers generate the best and second-best metric values, respectively. It is important to note that methods based on Shapley values, that apply a significant dimensionality reduction, exhibit a negligible performance loss in comparison with methods that do not include the recursive feature elimination step. Tables SM2, SM3, and SM4 in Supplemental material 2 show the performance metrics in percentage and the 95% CIs yielded by the proposed detection methods for Canada, Japan, and South Africa, respectively.

**Table 2.** Performance metrics in percentage and the 95% confidence intervals (CIs) obtained by the proposed COVID-19 detection methods for Brazil and for 2020 and 2021.

| Year | Method | F<sub>1</sub>-scores | Specificity | Sensitivity | Precision |
| --- | --- | --- | --- | --- | --- |
| 2020 | RF | **84.24 (84.19 - 84.30)** | **82.88 (82.79 - 82.98)** | **83.43 (83.35 - 83.51)** | **85.08 (85.00 - 85.15)** |
|  | XGB | 80.56 (80.50 - 80.62) | 78.96 (78.86 - 79.06) | 79.59 (79.50 - 79.67) | 81.57 (81.49 - 81.65) |
|  | LGB | 80.28 (80.22 - 80.34) | 78.53 (78.43 - 78.63) | 79.37 (79.29 - 79.44) | 81.22 (81.14 - 81.30) |
|  | RF SHAP | 84.23 (84.18 - 84.28) | 82.87 (82.78 - 82.97) | 83.41 (83.33 - 83.49) | 85.07 (84.99 - 85.15) |
|  | XGB SHAP | 80.57 (80.51 - 80.63) | 78.97 (78.87 - 79.07) | 79.59 (79.51 - 79.66) | 81.58 (81.50 - 81.66) |
|  | LGB SHAP | 80.26 (80.21 - 80.32) | 78.53 (78.42 - 78.64) | 79.34 (79.26 - 79.41) | 81.21 (81.13 - 81.30) |
| 2021 | RF | **80.43 (80.39 - 80.47)** | **91.40 (91.37 - 91.43)** | **75.74 (75.68 - 75.80)** | **85.74 (85.68 - 85.79)** |
|  | XGB | 73.89 (73.85 - 73.94) | 88.75 (88.71 - 88.79) | 68.25 (68.19 - 68.31) | 80.55 (80.48 - 80.61) |
|  | LGB | 73.50 (73.45 - 73.54) | 88.54 (88.50 - 88.58) | 67.86 (67.80 - 67.91) | 80.16 (80.09 - 80.23) |
|  | RF SHAP | 79.11 (79.07 - 79.15) | 90.15 (90.11 - 90.18) | 74.90 (74.84 - 74.96) | 83.85 (83.79 - 83.91) |
|  | XGB SHAP | 72.53 (72.49 - 72.58) | 88.26 (88.22 - 88.30) | 66.69 (66.62 - 66.76) | 79.50 (79.43 - 79.57) |
|  | LGB SHAP | 73.49 (73.44 - 73.53) | 88.54 (88.50 - 88.58) | 67.84 (67.78 - 67.90) | 80.16 (80.10 - 80.22) |

Figure 2 presents the receiver operating characteristic (ROC) curves and 95% CIs produced by the implemented machine learning models for the four countries and 2020. More precisely, each ROC curve is derived by averaging ten realizations of the respective experiment, where different training and test sets are randomly generated at each trial. The training set contains 80% of the samples, while the test set contains the 20% remaining ones. Every ROC curve includes the area under the ROC curve (auROC) and its 95% CI. For 2020, the detection methods obtaining the best auROCs for each country are Brazil (RF: 0.884, 95% CI: 0.845 − 0.923), Canada (LGB_Shap: 0.913, 95% CI: 0.892 − 0.934), Japan (LGB: 0.880, 95% CI: 0.835 − 0.925), and South Africa (RF_Shap: 0.919, 95% CI: 0.871 − 0.967). Note that the lowest auROC value is obtained for Brazil (XGB_Shap: 0.853, 95% CI: 0.839 − 0.869).

**Figure 2:**
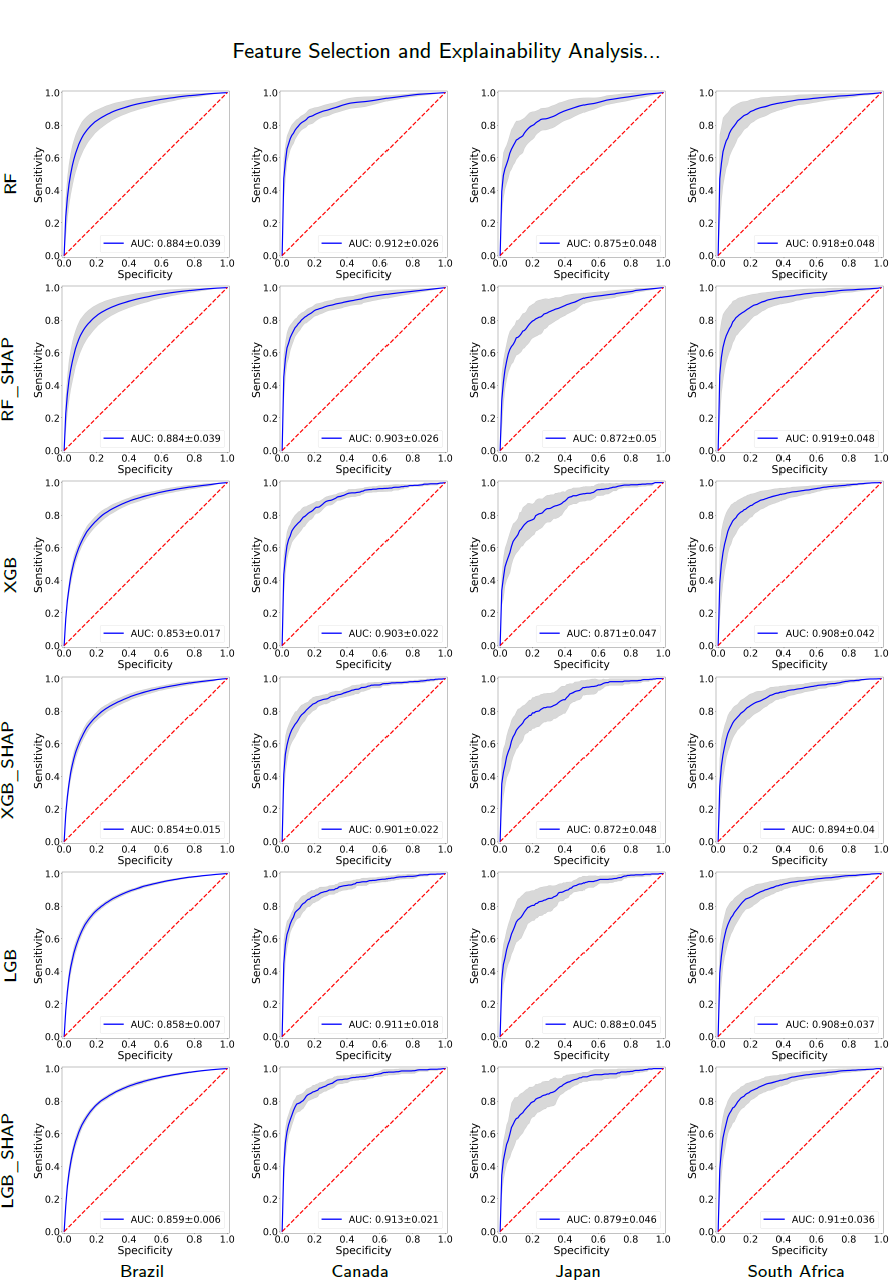
ROC curves and their 95% confidence intervals yielded by the proposed approach using different classification models in each of the four countries and for 2020. Each ROC curve includes the AUC corresponding value.

On the other hand, Figure 3 illustrates the ROC curves and their 95% CIs obtained by the proposed COVID-19 detection models for the four countries and 2021. The detection methods yielding the best auROC values for every country are Brazil (RF: 0.879, 95% CI: 0.805 − 0.953), Canada (XGB: 0.903, 95% CI: 0.889 − 0.917), Japan (RF: 0.918, 95% CI: 0.881 − 0.955), and South Africa (RF: 0.887, 95% CI: 0.836 − 0.938). For 2021, the lowest auROC value is also obtained for Brazil (LGB: 0.817, 95% CI: 0.716 − 0.918).

**Figure 3:**
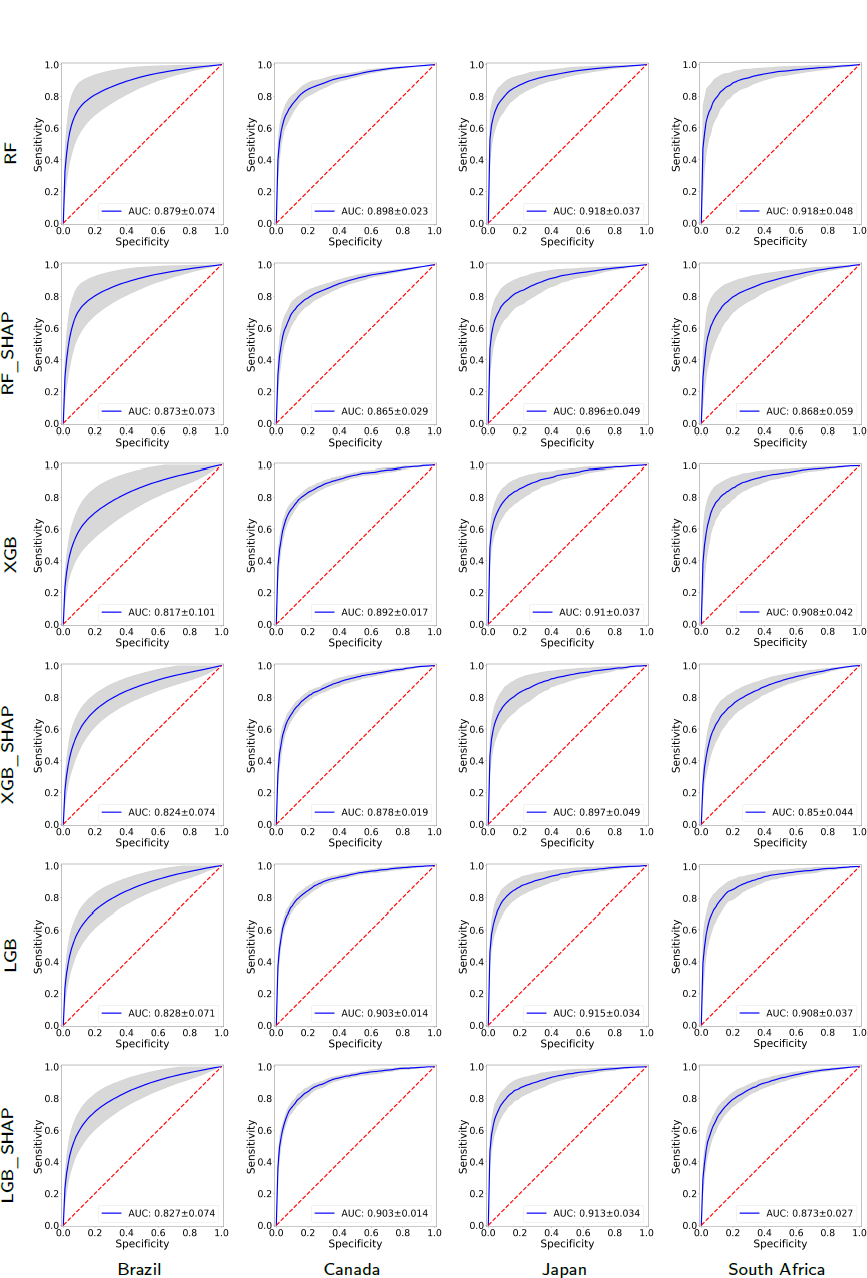
ROC curves and their 95% confidence intervals yielded by the proposed approach using different classification models in each of the four countries and for 2021. Each ROC curve includes the AUC corresponding value.

Figure 4 displays the F_1_-scores and the corresponding 95% CIs obtained by different COVID-19 detection methods for the four countries and for 2020 and 2021. Notice that F_1_-scores are presented in descending order to identify the best performance. Particularly, we display the F_1_-scores produced by the detection methods based on RF, XGB, LGB, RF_Shap, XGB_Shap, and LGB_Shap. For comparison purposes, we also included the F_1_-scores obtained by previously reported detection techniques such as Menni [7], Smith [6], Shoer [17], Mika [19], and Astley [2]. In Supplemental Material 2, Table SM1 shows the numerical values of the F_1_-scores and the 95% CIs obtained by the different detection methods for the four countries and for 2020 and 2021. Specifically, for 2020, the detection methods that yield the best F_1_-scores for every country are: Brazil (RF: 84.24%, 95% CI: 84.19% to 84.29%), Canada (XGB: 62.53%, 95% CI: 61.98% to 63.09%), Japan (XGB_Shap: 59.70%, 95% CI: 58.82% to 60.57%), and South Africa (RF_Shap: 81.88%, 95% CI: 81.68% to 82.09%). In contrast, the approaches generating the lowest F_1_-scores for each country are: Brazil (Menni: 65.56%, 95% CI: 65.48% to 65.54%), Canada (Astley: 48.29%, 95% CI: 47.58% to 49.00%), Japan (Astley: 44.13%, 95% CI: 43.32% to 44.93%), and South Africa (Menni: 61.39%, 95% CI: 61.07% to 61.70%). For 2021, the methods yielding the best F_1_-scores for every country are: Brazil (RF: 80.43%, 95% CI: 80.39% to 80.47%), Canada (RF: 63.80%, 95% CI: 63.52% to 64.08%), Japan (RF: 70.11%, 95% CI: 69.84% to 70.37%), and South Africa (RF: 77.69%, 95% CI: 77.53% to 77.85%). On the contrary, the detection methods producing the lowest F_1_-scores for every country are: Brazil (Menni: 59.24%, 95% CI: 59.18% to 59.31%), Canada (Shoer: 41.10%, 95% CI: 40.84% to 41.36%), Japan (Shoer: 45.42%, 95% CI: 45.07% to 45.78%), and South Africa (Menni: 58.28%, 95% CI: 58.06% to 58.50%). It is worth noting that the proposed COVID-19 detection methods outperform previously reported techniques for the four countries and for the two periods under test. F_1_ scores for 2020 and 2021 are presented in Figure SM1 in the Supplemental Material to compare the performance of each detection method across the countries under test. As shown in this figure, each method generates the best F_1_ scores for Brazil or South Africa. Table 1 highlights that these countries exhibit TPR values that are at least three times larger than those of Canada and Japan.

**Figure 4:**
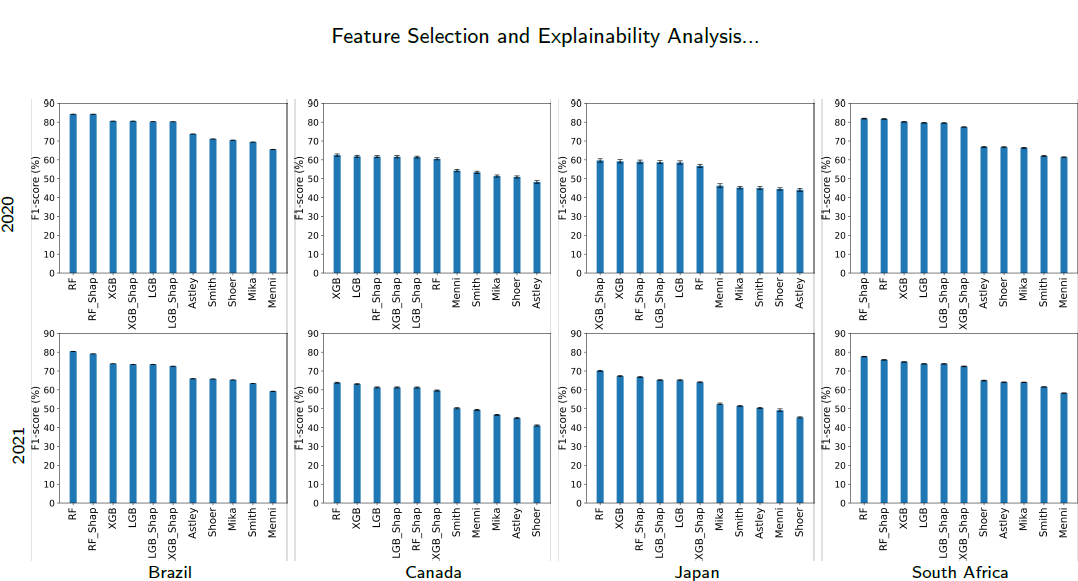
F_1_-scores and the 95% CIs yielded by various COVID-19 detection methods for the four countries and for 2020 and 2021.

Finally, to evaluate the proposed COVID-19 detection methods in a practical problem, we estimate the normalized COVID-19 daily case curves for the four countries from January 1, 2021, to June 25, 2022. Using the proposed detection methods based on RF, LGB, and XGB, we first identify the daily COVID-19 active cases in the interval of interest. Afterward, the estimated daily case curves are normalized with respect to the maximum value. A comparison of the daily case curves obtained by the proposed detection methods for the four countries is shown in Figure 5. In addition, we include the normalized COVID-19 daily case curve provided by the respective national healthcare system for comparison. In each country, we present a Pearson correlation coefficient between the curve obtained by each proposed detection methodology and the curve provided by the national healthcare system. The best correlation coefficients are generated by the proposed approach based on the LGB classifier, i.e., Brazil (LGB: 0.94), Canada (LGB: 0.59), Japan (LGB: 0.98), and South Africa (LGB: 0.88). As an important note, the daily case curves generated by these detection methods have been used by the CoronaSurveys Project (https://coronasurveys.org), a collaboration between several academic institutions aimed at providing global pandemic surveillance based on surveys, to estimate daily active cases for more than 150 countries/territories [32].

**Figure 5:**
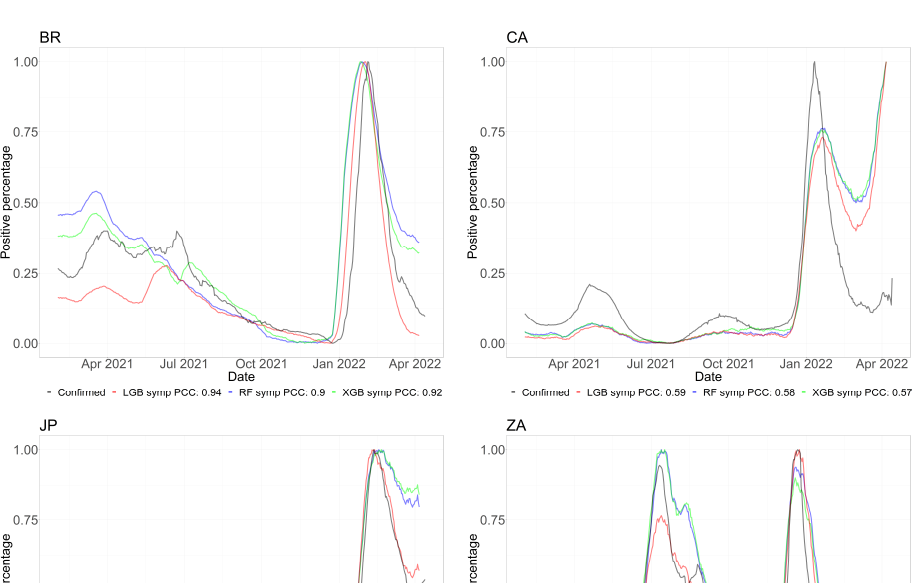
Normalized daily case curves generated by the proposed detection methods based on RF, LGB, and XGB. Normalized daily case curves provided by the respective healthcare system is also displayed for each country.

### 3.2. Explainability analysis

This work considers a local and post-hoc approach based on Shapley values and a global ante-hoc approach based on the RF because of its superior accuracy with respect to the other tree-based methods considered in this work.

#### 3.2.1. Explainability analysis using Shapley values

We now proceed to analyze the explainability based on the results given by the different techniques, using (or not) the Shapley-based feature removal, to estimate positive cases, for the four countries and for 2020 and 2021 (see Figures 6, 7). We will delve deeper into the model that gave the best results, both in its full version and with the Shapley-based feature reduction. In the analysis, we have considered that the variables with Shapley values greater than 0.05 are relevant to consider, as stipulated in the literature [21, 31].

- Year 2020: According to Figure 6, in the case of RF the variable B1.10.1 is relevant in all cases (countries), while in other cases, the variable B1.10.2 is also relevant as C0.2.1 (it is the most relevant for Japan but with a low relevance value of less than 0.03). This makes sense because these variables refer to the loss of smell/taste (B1.10.1 and B1.10.2), and the fact of having gone to a market, store or pharmacy (C0.2.1). By 2020, the most widespread symptom was loss of smell and taste. B1.10.1, even in the other methods, had an even higher relevance value, reaching values greater than 0.1 in LGB and LGB.SHAP for Brazil and South Africa, and 0.08 in XGB and XGB.SHAP. Also, variable B1bx10.1, which means that a usual symptom was the loss of taste or smell, appears relevant in the case of South Africa for RF (close to 0.03). This is very much in agreement with what happened for the variant that prevailed in 2020.

**Figure 6:**
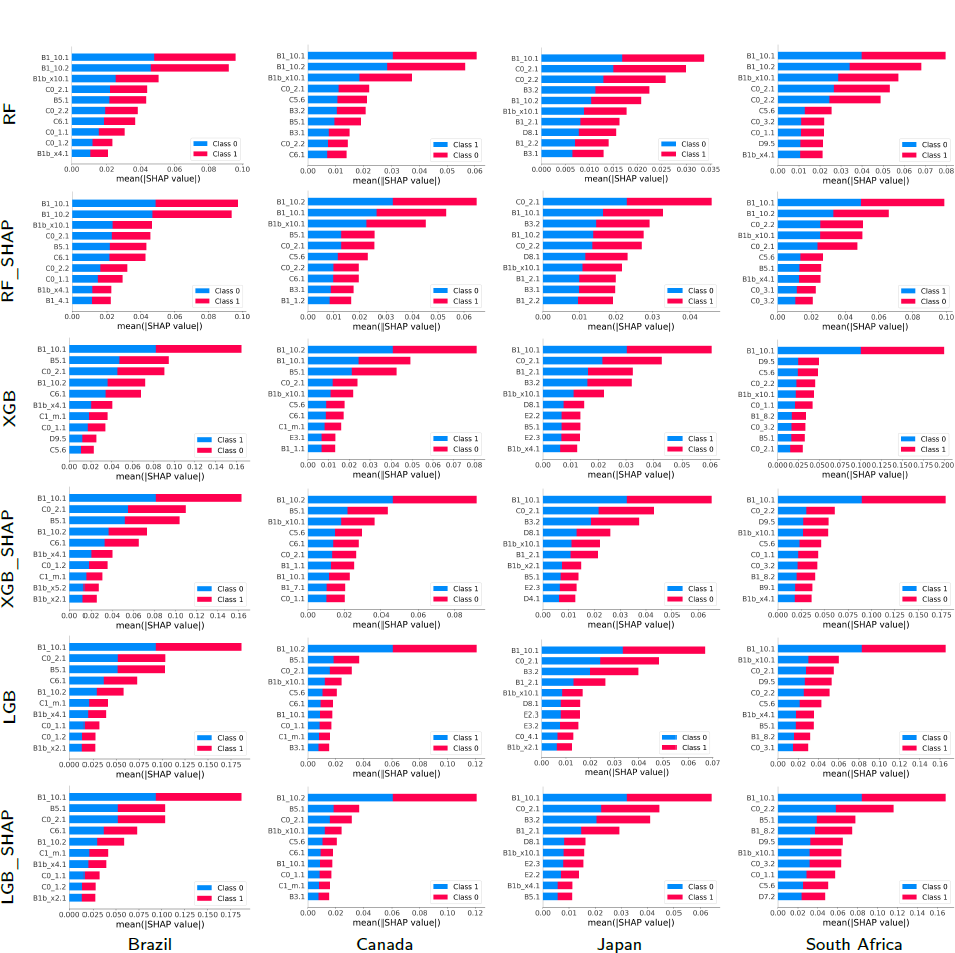
SHAP values and their impact on the detection output of the 10 most relevant features obtained by the proposed approach using different classification models for the four countries and for 2020.

**Figure 7:**
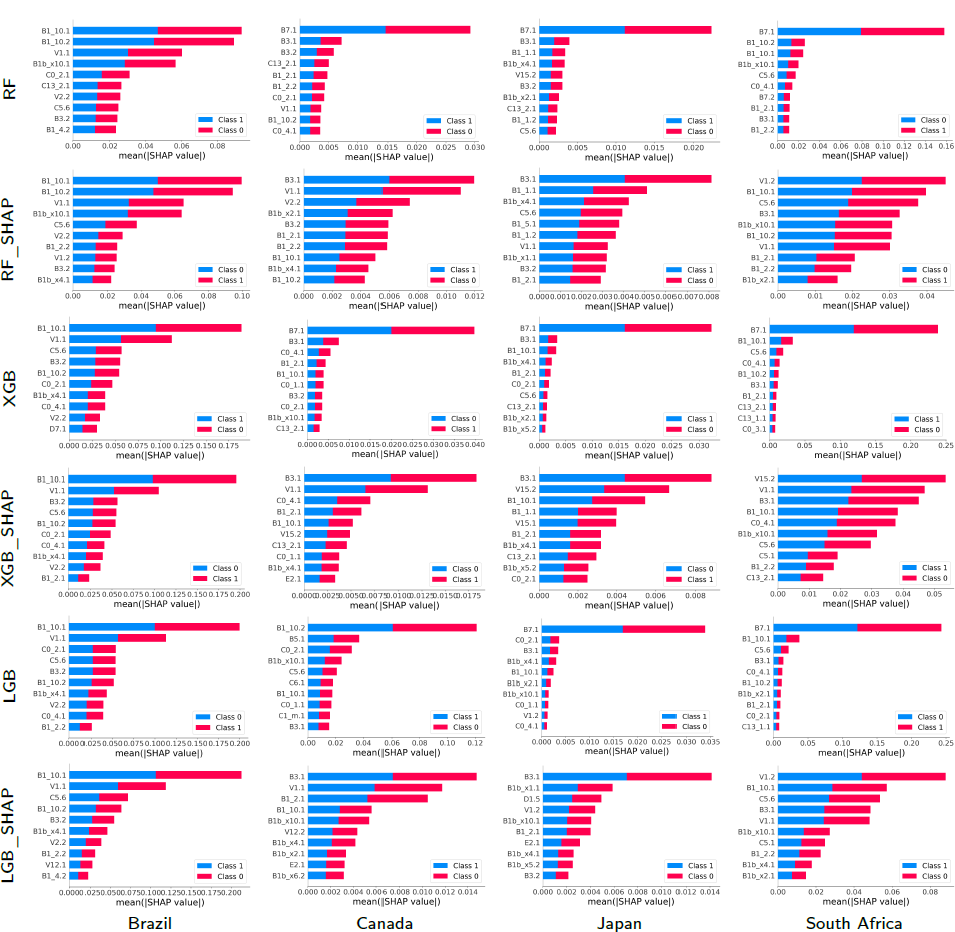
SHAP values and their impact on the detection output of the 10 most relevant features obtained by the proposed approach using different classification models for the four countries and for 2021.

In general, for 2020, the most important variable in every model is the loss of smell or taste. Similarly to what was determined by [33, 34], this was the most representative symptom of COVID-19 with the first variant, the only one present in 2020. Along with the variables described in the previous paragraph, other variables somewhat relevant (in some cases, with Shapley values greater than 0.04) were B5.1 (spent time with COVID-19 infected people), C6.1 (Not spend time with someone outside your household) or C5.6 (Not going out for a week) to determine if an individual is COVID-19 positive.

To continue with the explainability analysis, and as an example, we did our own ranking to determine the relevance of the features in general, according to the order using the Shapley value in which a feature appears in each technique for each country. For this ranking, the first 10 variables were considered according to the Shapley value for each technique in each country, and a 10 was assigned to the one with the highest Shapley value, 9 to the next, and so on. Then, at the end, the values obtained by each feature in each country-technique pair are added to obtain its position in our ranking.

Table 3 lists only the first 10 variables according to that ranking. We can see that the variable B1.10.1 has a value much higher than the rest (218), and with C0.2.1 (192) they are very far from the rest. They are the same variables that we had determined before as the most relevant, which corroborates our previous explainability analysis. B1.10.1 has the highest value in almost all cases, and only in Canada, for some techniques, it has a low value (for example, XGB with 3). This happens less in C0.2.1, since its lowest value is 5 (also for XGB and Canada). The rest of the techniques have at least once a value of 0, which means that they are not among the first 10 variables according to the Shapley value in that technique and country.

**Table 3.**
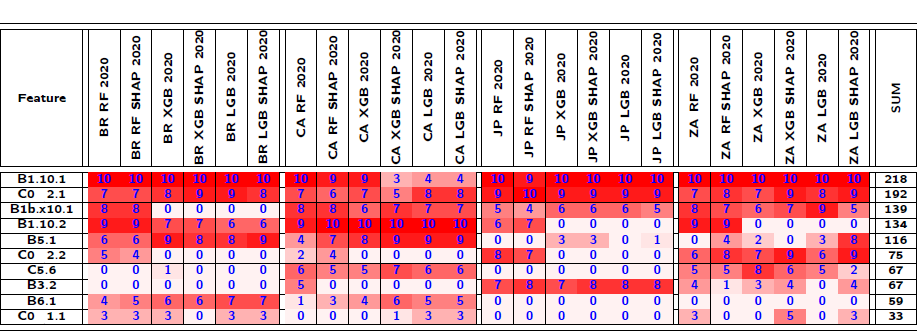
Ranking of the features based on the Shapley values for the entire set of countries for 2020.

We also see some cases where the techniques in some countries only use a few of the best ranked variables according to our ranking. For example, the case of Brazil with XGB using Shapley, or Japan with XGB, which use only 5 of the first 10 variables of our ranking. That implies that they have other more relevant variables, in addition to those 5, to the first 10 established by our ranking (for example, V1.1 has the second largest Shapley value in Japan with XGB). We also see that there is no technique for any country that uses those 10 best ranked variables according to our criteria, only RF for Canada used 9 of them.

- Year 2021: This year was characterized by the appearance of several variants of COVID-19 (Delta, Omicron, etc.) and by the massive vaccination of the population against COVID-19. In this case, the variable B1.10.1 continued to appear as one of the most relevant variables, and in some cases B1.10.2. However, other variables that also appeared with great relevance were V1.1 and B7.1. These variables refer to whether the person has been vaccinated against COVID-19 (V1.1), and if in the last 4 weeks the person did any paid work (B7.1). Particularly, V1.1 is relevant in Brazil (despite being a country where some government instances promoted the denial of the positive effect of vaccination), and B7.1 is the most relevant on several occasions (for example, for RF and XGB in Canada, South Africa and Japan, although with low relevance values, of the order of 0.015). Other variables that appear with some relevance in some cases are B3.1 that deals with whether someone in the local community is known to have been ill with fever, cough, or difficulty breathing (for example, the most relevant for RF.SHAP in Canada and Japan, among others, but with a very low relevance (less than 0.004)), B15.2 which is if the person has had an appointment to receive a COVID-19 vaccine (the most relevant for XGB.SHAP in South Africa), and V1.2 which deals with whether to have a COVID-19 vaccine (the most relevant for XGB.SHAP in South Africa with 0.05). Variable B3.1 is the most relevant for all SHAP-based methods for Canada and Japan, which indicates whether anyone in the local community was known to have fever and cough or shortness of breath as one of the causes of seropositivity (rapid spread of the virus). Similarly, variable B7.1 is the most relevant for all methods without SHAP for Japan, Canada and South Africa (contagion from going to work, also linked to the rapid spread of the virus). Finally, V1.2 is the most relevant for all methods with SHAP in South Africa, indicating the fact of not having been vaccinated as one of the reasons for high seropositivity in that country. In the case of Brazil, the most relevant variables were again B1.10.1, B1.10.2 and V1.1. Also to note that C0.2.1 ceased to be relevant in 2021. For 2021, again, one of the most important features is the loss of smell or taste. However, with less relevance in some countries as it was in 2020 due to the COVID-19 variants during this year along with the vaccination. In addition, variables linked to facilitating the spread (such as going to work (B7.1) or if the person had acquaintances with symptoms (B3.1)) appear as reasons for seropositivity, or the case of not having been vaccinated yet (South Africa and V1.2). Thus, 2021 has some of the same important features as 2020 but the variables related to the vaccines and the rapid spread of the virus also play a key role.

#### 3.2.2. Explainability analysis using the range of features given by RF

In this part, we carry out an explainability analysis for RF becasue it is the technique that showed the best quality (see Figures 8 and 9). With RF, various measures of feature importance can be used for an explainability analysis. In this work, we have used MDA for being one of the best feature-importance measures according to the literature [26, 30].

- Year 2020: It is interesting to see that again, the most relevant variables are B1.10.1, B1.10.2, as well as B1bx10.1. The relevance order changes in some cases, as for RF (B1.10.1 is the variable that appears as relevant most frequently for RF using the Shapley values, and in this case is shared with B1.10.2). In general, the relevance values of the most relevant variables are always high, and occasionally very high (for example, B1.10.1 for Canada close to 0.8).

**Figure 8:**
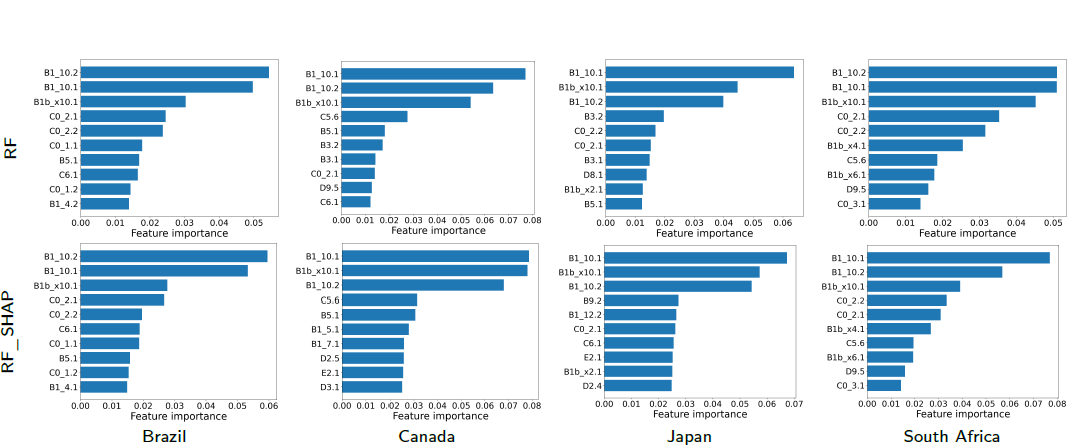
Feature importance of the 10 most relevant input variables obtained by classification models based on the random forest method for the four countries and for 2020.

**Figure 9:**
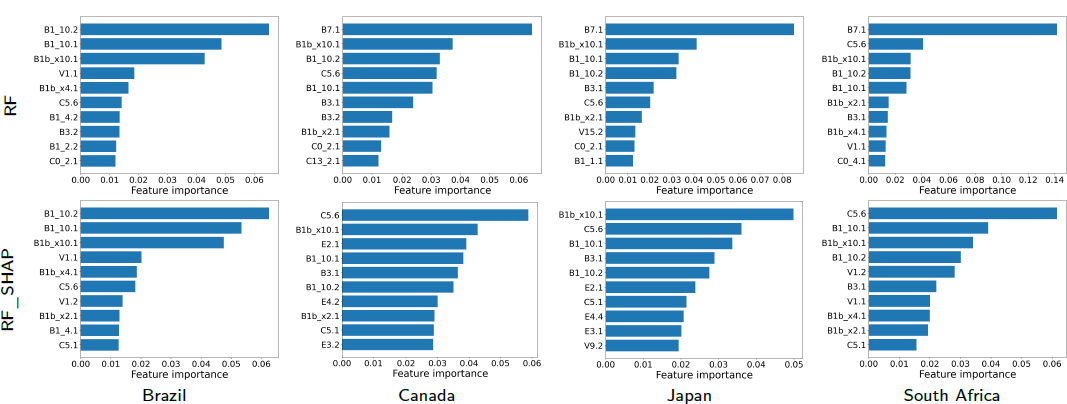
Feature importance of the 10 most relevant input variables obtained by classification models based on the random forest method for the four countries and for 2021. Another aspect to note is how in some cases the most frequent variables change, as in the case of Japan with C0.2.1 (it was among the most relevant variables in the Shapley values and was no longer among the three most relevant in the RF ranking). Finally, B1.10.1 and B1.10.2 are always the most relevant, with values greater than 0.05 regardless of whether RF or RF.SHAP is used. This clearly indicates that the variables linked to loss of smell/taste are the fundamental ones for estimating seropositivity to COVID-19 in the case of RF and RF.SHAP. Year 2021: In the case of Brazil, the most relevant variables were once again B1.10.1, B1.10.2, with B1.10.2 now being more relevant. On the other hand, the variable V1.1 disappears from the relevant group (which makes sense, because Brazil was a country where the denial of the positive effect of vaccination was promoted from some of the government instances). Also, variable B7.1 continues to be the most relevant for all methods without SHAP for Japan, Canada and South Africa, being very decisive in Japan and Canada (values greater than 0.08, and the next with relevance values around 0.04). Another variable that is no longer relevant is B3.1, with two variables appearing as highly relevant, B1bx10.1 and C5.6, particularly for SHAP-based techniques. B1bx10.1 is relevant in Japan, which means that a common symptom was the loss of taste or smell; and C5.6 in South Africa and Canada, and it has to do with not having been in public during the last 7 days (self-care of people).

## 4. Discussion

This study presents a machine-learning approach to detecting COVID-19-active cases based on three classification models: random forest (RF), light gradient boosting (LGB), and extreme gradient boosting (XGB). More precisely, the proposed detection approach predicts active cases using the entire set of variables collected from the UMD-CTIS questionnaires. These questionnaires record a wide range of individual features such as gender, age group, vaccination acceptance, and isolation measures. In addition, we introduce a feature-reduction approach that uses a recursive feature-elimination (RFE) strategy to train the classification model. A key objective of RFE is to identify and keep relevant features based on Shapley values without compromising detection accuracy. It is pertinent to mention that the proposed method is evaluated on UMD-CTIS data extracted from four countries: Brazil, Canada, Japan, and South Africa, and two periods: 2020 and 2021. Specifically, we consider experiments with at least one symptom reported within the past 24 hours and a test result within the past 14 days. Extracted datasets may contain biases, limitations, and missing values. In countries where demographic information can be a significant factor in detecting active cases, such as isolated communities and islands, the data may be affected by biases due to the homogenization of the population. In addition, biases can arise from assuming that all populations have Internet access uniformly. To reduce biases, we randomly select a limited set of experiments as the training set to optimize the classification models.

The proposed approach has shown competitive performance for the four countries and 2020 and 2021. In particular, the feature selection stage removes a large number of irrelevant variables with a negligible impact on classification accuracy. According to different quality metrics (such as F_1_-score, sensitivity, specificity, precision, and AUC), RF and RF SHAP models exhibit the most accurate detection performances across the four countries and for 2020 and 2021. We also compare the performance of the developed technique to those yielded by previously reported approaches based on surveys. The proposed detection methods outperform the state-of-the-art methods for the four countries in terms of F_1_-score. The RF-based approach obtained the highest results, regardless of whether or not feature selection was used. In the final step, we use the developed detection approach to construct the normalized daily-case curve for the four countries between January 1, 2021, and June 25, 2022, to observe pandemic trends. In comparison with official records, these estimated curves provide consistent tracking of pandemic evolution. Therefore, taking into account both the proposed detection approaches and the massive amount of data provided by the UMD-CTIS questionnaires, we can reliably track pandemic indicator trends in a similar way to that provided by public healthcare systems.

Regarding the explainability analysis, the variables that appear most frequently are the loss of smell or taste (B1.10.1/B1.10.2), regardless of the year, country, the technique used for the explainability analysis, the prediction technique, or dataset reduced or not by the Shapley method. Other variables appear in some specific cases (countries, forecasting techniques, etc.). For example, using the Shapley method for explainability analysis in 2020, the variable C0.2.1 (the fact of having gone to a market, store or pharmacy) is relevant in some countries. Also, in 2021, the variable V1.1 is relevant in Brazil (whether the person has been vaccinated against COVID-19), the variable B7.1 is relevant in some countries (if in the last 4 weeks the person did any paid work) for the cases without data reduction, and B3.1 (it indicates whether anyone in the local community was known to have fever and cough) for cases with data reduction using the Shapley method. The same happens using the RF ranking for the explainability analysis: new variables appear (such as C5.6) or some existing ones disappear (such as C0.2.1 and B3.1).

In any case, attribute-based explainability analysis shows the relevant variables for decision makers to detect seropositivity very quickly. This is valid both for the case of using the ranking given by RF or the Shapley values. However, it is important to highlight that although they present common variables, between the two techniques there are some differences between those that are considered to be more relevant. For example, V1.1 appears as relevant in the Shapley method for Brazil and disappears in the RF ranking, which makes more sense because in Brazil, the vaccination campaign did not have a strong support from the government.

Thus, RF seems sufficient to achieve good results and explain the results obtained (explainability analysis). But although explainability is aimed at the understanding by experts and non-experts, there are no designs or formal evaluations on the human usability of the methods analyzed in this work. This is pending work, which goes beyond simple representations of explanations. At the same time, the analysis of the variables by classes has rarely been carried out (in the case of Shapley, the values by class are similar/symmetrical), which opens up a space for research for the development of techniques that allow analyzing the possibility of explainability by classes (which characteristics/variables are relevant for each class).

## 5. Declarations

### 5.1. Ethical Declaration

The Ethics Board (IRB) of IMDEA Networks Institute gave ethical approval for this work on 2021/07/05. IMDEA Networks has signed Data Use Agreements with Facebook, Carnegie Mellon University (CMU) and the University of Maryland (UMD) to access their data, specifically UMD project 1587016-3 entitled C-SPEC: Symptom Survey: COVID-19 and CMU project STUDY2020_00000162 entitled ILI Community-Surveillance Study. The data used in this study was collected by the University of Maryland through The University of Maryland Social Data Science Center Global COVID-19 Trends and Impact Survey in partnership with Facebook. Informed consent has been obtained from all participants in this survey by this institution. All the methods in this study have been carried out in accordance with relevant of ethics and privacy guidelines and regulations.

### 5.2. Availability of Data and Materials

The data presented in this paper (in aggregated form) and the programs used to process it will be openly accessible at https://github.com/GCGImdea/coronasurveys/. The microdata of the CTIS survey from which the aggregated data was obtained cannot be shared, as per the Data Use Agreements signed with Facebook, Carnegie Mellon University (CMU) and the University of Maryland (UMD).

## Data Availability

https://github.com/GCGImdea/coronasurveys/

## Supplemental Materials: Feature Selection for an Explainability Analysis in Detection of COVID-19 Active Cases in Countries from Facebook User-Based Online Surveys

### 1. Questions of the survey

- **B1 In the last 24 hours, have you had any of the following?** Fever (B1_1), Cough (B1_2), Difficulty breathing (B1_3), Fatigue (B1_4), Stuffy or runny nose (B1_5), Aches or muscle pain (B1_6), Sore throat (B1_7), Chest pain (B1_8), Nausea (B1_9), Loss of smell or taste (B1_10), Headache (B1_12), Chills (B1_13).
- **B1b Are any of these symptoms unusual for you?** Fever (B1b_x1), Cough (B1b_x2), Difficulty breathing (B1b_x3), Fatigue (Bb1_x4), Stuffy or runny nose (B1b_x5), Aches or muscle pain (B1b_x6), Sore throat (B1b_x7), Chest pain (B1b_x8), Nausea (B1b_x9), Loss of smell or taste (B1b_x10), Headache (B1b_x12), Chills (B1b_x13).
- **B3 Do you personally know anyone in your local community who is sick with a fever and either a cough or difficulty breathing?** Yes(1), No(2).
- **B5 Have you spent time with any of these people in the last 7 days?** Yes(1), No(2)^1^
- **B6 Have you ever been tested for coronavirus (COVID-19)?** Yes(1), No(2)^1^
- **B7 Have you been tested for COVID-19 in the past 14 days?** Yes(1), No(2).
- **B8 Did your most recent test find that you had COVID-19?** Yes(1), No(2), I don’t know(3).
- **B9 Did you have to pay anything out-of-pocket for this test?** Yes(1), No(2), I don’t know(3).^1^
- **B10 Have you or your household had to reduce spending on things you need (such as food,housing, or medication) because of the cost you paid to get the coronavirus (COVID-19) test?** Yes(1), No(2), I don’t know(3).^1^
- **B10 Have you or your household had to reduce spending on things you need (such as food, housing, or medication) because of the cost you paid to get the coronavirus (COVID-19) test?** Yes(1), No(2), I don’t know(3).^1^
- **B12_1 Do any of the following reasons describe why you haven’t been tested for coronavirus (COVID-19) in the last 14 days? I tried to get a test but was not able to get one** Yes(1), No(2).^1^
- **B12_2 Do any of the following reasons describe why you haven’t been tested for coronavirus (COVID-19) in the last 14 days? I don’t know where to go** Yes(1), No(2).^1^
- **B12_3 Do any of the following reasons describe why you haven’t been tested for coronavirus (COVID-19) in the last 14 days? I can’t afford the cost of the test** Yes(1), No(2).^1^
- **B12_4 Do any of the following reasons describe why you haven’t been tested for coronavirus (COVID-19) in the last 14 days? I don’t have time to get tested** Yes(1), No(2).^1^
- **B12_5 Do any of the following reasons describe why you haven’t been tested for coronavirus (COVID-19) in the last 14 days? I am unable to travel to a testing location (including because of transportation cost, safety, or physical limitations)** Yes(1), No(2).^1^
- **B12_6 Do any of the following reasons describe why you haven’t been tested for coronavirus (COVID-19) in the last 14 days? I am worried about bad things happening to me or my family (including discrimination, government policies, and social stigma)** Yes(1), No(2).^1^
- **B13_1 In the last 30 days, was there any time when you needed any of the following health services or products but could not get it? Emergency transportation services or emergency rescue** Yes(1), No(2).^1^
- **B13_2 In the last 30 days, was there any time when you needed any of the following health services or products but could not get it? Medical care with overnight stay in any type of facility** Yes(1), No(2).^1^
- **B13_3 In the last 30 days, was there any time when you needed any of the following health services or products but could not get it? Medical or dental care or treatment without an overnight stay** Yes(1), No(2).^1^
- **B13_4 In the last 30 days, was there any time when you needed any of the following health services or products but could not get it? Preventive health services (including immunization/vaccination, family planning, prenatal/postnatal care, routine check-up services)** Yes(1), No(2).^1^
- **B13_5 In the last 30 days, was there any time when you needed any of the following health services or products but could not get it? Medication** Yes(1), No(2).^1^
- **B13_6 In the last 30 days, was there any time when you needed any of the following health services or products but could not get it? Mask, medical gloves, or other protective equipment** Yes(1), No(2).^1^
- **B13_7 In the last 30 days, was there any time when you needed any of the following health services or products but could not get it? Eyeglasses, hearing aid, crutches, band-aids/plasters, thermometer, or any other health product** Yes(1), No(2).^1^
- **B14_1 In the last 30 days, were you unable to get needed treatment, services, medicine, or medical products for any of the following reasons? I didn’t know where to go** Yes(1), No(2).^1^
- **B14_2 In the last 30 days, were you unable to get needed treatment, services, medicine, or medical products for any of the following reasons? I couldn’t afford the treatment, service, or product** Yes(1), No(2).^1^
- **B14_3 In the last 30 days, were you unable to get needed treatment, services, medicine, or medical products for any of the following reasons? I was unable to travel to the health care provider (including because of transportation cost, safety, or physical limitations)** Yes(1), No(2).^1^
- **B14_4 In the last 30 days, were you unable to get needed treatment, services, medicine, or medical products for any of the following reasons? I was afraid of being infected at the health care provider** Yes(1), No(2).^1^
- **B14_5 In the last 30 days, were you unable to get needed treatment, services, medicine, or medical products for any of the following reasons?The treatment, service, or product was not available** Yes(1), No(2).^1^
- **C0_1 In the last 24 hours, have you done any of the following? Gone to work outside the place where you are currently staying** Yes(1), No(2)
- **C0_2 In the last 24 hours, have you done any of the following? Gone to a market, grocery store, or pharmacy** Yes(1), No(2)
- **C0_3 In the last 24 hours, have you done any of the following? Gone to a restaurant, cafe, or shopping center** Yes(1), No(2)
- **C0_4 In the last 24 hours, have you done any of the following? Spent time with someone who isn’t currently staying with you** Yes(1), No(2)
- **C0_5 In the last 24 hours, have you done any of the following? Attended a public event with more than 10 people** Yes(1), No(2)
- **C0_6 In the last 24 hours, have you done any of the following? Used public transit** Yes(1), No(2)
- **C1_m In the last 24 hours, have you had direct contact with anyone who is not staying with you? Direct contact means spending longer than one minute within two meters of someone or touching, including shaking hands, hugging, or kissing.** Yes(1), No(2)^1^
- **C2 How many people, who are not staying with you, have you had this kind of direct contact with in the last 24 hours?** 1-4 people(1), 5-9 people(2), 10-19 people (3), 20 or more(4).^1^
- **C3 Do you have access to soap and water for washing your hands at the place where you are currently staying?** Yes(1), No(2)^1^
- **C5 In the last 7 days, how often did you wear a mask when in public?** All of the time(1), Most of the time(2), Some of the time(3), A little of the time(4), None of the time(5), I have not been in public during the last 7 days(6)
- **C6 In the last 7 days, how many days have you spent time with people who aren’t staying with you?** 0 days(1), 1 day(2), 2-4 days(3),5-7 days(4).^1^
- **C7 In the last 24 hours, about how many times have you washed your hands with soap and water or used hand sanitizer?** 0 times (1), 1-2 times (2), 3-6 times (3),7 or more times (4).^1^
- **C8 Do you have access to soap and water for washing your hands at the place where you are currently staying?**Yes (1), No (2)^1^
- **C13a During which activities in the past 24 hours did you wear a mask? Please select all that apply.** Gone to work or school indoors, outside the place where you are currently staying (1), Gone to an indoor market, grocery store, or pharmacy (2), Had a drink or meal indoors at a bar, restaurant, or cafe (3), Spent time indoors with someone who isn’t currently staying with you (4), Attended an indoor event with more than 10 people (5),Used public transit (6). ^2^
- **D1 During the last 7 days, how often did you feel so nervous that nothing could calm you down?** All of the time(1), Most of the time(2), Some of the time(3), A little of the time(4), None of the time(5).
- **D2 During the last 7 days, how often did you feel so depressed that nothing could cheer you up?** All of the time(1), Most of the time(2), Some of the time(3), A little of the time(4), None of the time(5).
- **D3 How worried are you that you or someone in your immediate family might become seriously ill from coronavirus (COVID-19)?** Very worried(1), Somewhat worried(2), Not too worried(3), Not worried at all(4).^1^
- **D4 How worried are you about having enough to eat in the next week?** Very worried(1), Somewhat worried(2), Not too worried(3), Not worried at all(4).
- **D5 How worried are you about your household’s finances in the next month?** Very worried(1), Somewhat worried(2), Not too worried(3), Not worried at all(4).
- **D7 In the past 4 weeks, did you do any work for pay? By work for pay, we mean any kind of business, farming, or other activity to earn money, even if only for one hour.** Yes (1), No(2).
- **D8 Before February 2020, were you working for pay, or doing any kind of business, farming, or other activity to earn money?** Yes(1), No(2)^1^
- **D9 Why did you stop working?** My employer closed for coronavirus-related reasons (1), My employer closed for another reason (2), I was laid off or furloughed (3), I am a seasonal worker (4), I was ill or quarantined (5), I needed to care for someone (6), Other (7)^1^
- **D10 What is the main activity of the business or organization in which you work?**Agriculture (1),Buying and selling (2), Construction (3),Education (4),Electricity / water / gas / waste (5),Financial / insurance / real estate services (6), Health (7), Manufacturing (8), Mining (9), Personal services (10), Professional / scientific / technical activities (11), Public administration (12), Tourism (13), Transportation (14), Other (15).
- **E2 Which of these best describes the area where you are currently staying?** City(1), Town(2), Village or rural area(3).
- **E3 What is your gender?** Male(1), Female(2), Other(3), Prefer not to answer(4)
- **E4 What is your age?** 18-24 years(1), 25-34 years(2), 35-44 years(3), 45-54 years(4), 55-64 years(5), 65-74 years(6), 75 years or older(7).
- **V1 Have you had a COVID-19 vaccination?**Yes (1), No (2),I don’t know (3)^2^
- **V2 How many COVID-19 vaccinations have you received?**1 vaccination or dose (1), 2 vaccinations or doses (2),I don’t know (3)^2^
- **V3 If a vaccine to prevent COVID-19 were offered to you today, would you choose to get vaccinated?** Yes, definitely (1), Yes, probably (2), No, probably not (3), No, definitely not (4)^2^
- **V5a Which of the following, if any, are reasons that you definitely wouldn’t choose to get a COVID-19 vaccine? Please select all that apply.** I am concerned about possible side effects of a COVID-19 vaccine (1), I don’t know if a COVID-19 vaccine will work (2), I don’t believe I need a COVID-19 vaccine (3),I don’t like vaccines (4), I plan to wait and see if it is safe and may get it later (5), I think other people need it more than I do right now (6), I am concerned about the cost of a COVID-19 vaccine (7), It is against my religious beliefs (8), I don’t trust the government (10), Other (9)^2^
- **V5b Which of the following, if any, are reasons that you probably wouldn’t choose to get a COVID-19 vaccine? Please select all that apply.** I am concerned about possible side effects of a COVID-19 vaccine (1), I don’t know if a COVID-19 vaccine will work (2), I don’t believe I need a COVID-19 vaccine (3), I don’t like vaccines (4), I plan to wait and see if it is safe and may get it later (5),I think other people need it more than I do right now (6), I am concerned about the cost of a COVID-19 vaccine (7), It is against my religious beliefs (8), I don’t trust the government (10), Other (9)^2^
- **V5c Which of the following, if any, are reasons that you probably wouldn’t choose to get a COVID-19 vaccine? Please select all that apply.** I am concerned about possible side effects of a COVID-19 vaccine (1), I don’t know if a COVID-19 vaccine will work (2), I don’t believe I need a COVID-19 vaccine (3), I don’t like vaccines (4), I plan to wait and see if it is safe and may get it later (5),I think other people need it more than I do right now (6), I am concerned about the cost of a COVID-19 vaccine (7), It is against my religious beliefs (8), I don’t trust the government (10), Other (9).^2^
- **V6 Why don’t you believe that you need a COVID-19 vaccine? Please select all that apply.**I already had COVID-19 (1), I do not spend time with any high-risk people (2), I am not a member of a high-risk group (3), I plan to use masks or other precautions instead (4), I don’t believe COVID-19 is a serious illness (5), I don’t think vaccines are beneficial (6), Other (7).^2^
- **V10 Have you ever been told by a doctor, nurse, or other health professional that you have any of the following medical conditions? Please select all that apply.** Asthma (1), Chronic lung disease such as COPD, chronic bronchitis, or emphysema (2), Cancer (3), Diabetes (4), High blood pressure (5), Kidney disease (6), Weakened or compromised immune system (7), Heart attack, heart disease, or other heart condition (8), Obesity (9), one of these (10).^2^
- **V15 Do you have an appointment to receive a COVID-19 vaccine?** Yes (1),No (2)^2^
- **V16 Have you tried to get an appointment to receive a COVID-19 vaccine?** Yes (1),No (2)^2^

### 2. Results

**Figure SM1:**
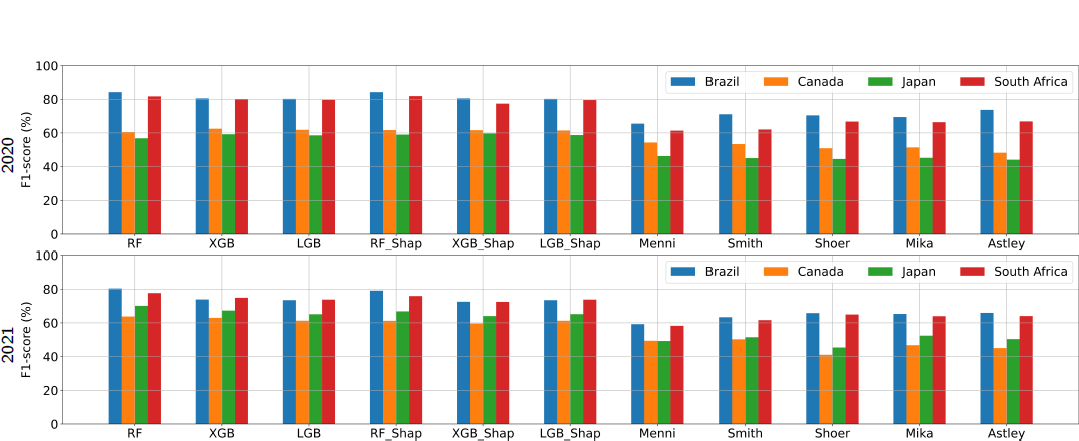
F_1_-scores and the 95% CIs for 2020 and 2021 generated by the various COVID-19 detection methods.

**Table SM1.**
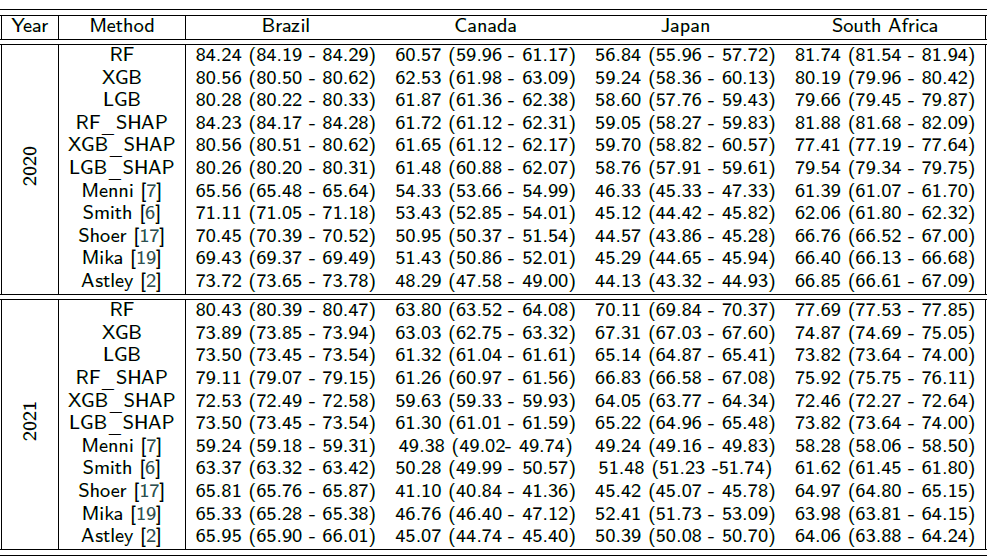
F_1_-scores and the 95% CIs yielded by various COVID-19 detection methods for the four countries and for 2020 and 2021.

**Table SM2.**
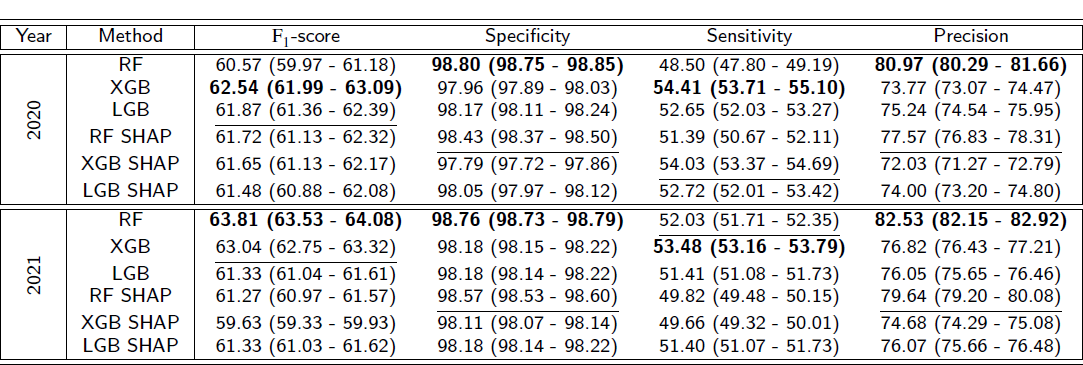
Performance metrics in % and the 95% CIs obtained by the proposed COVID-19 detection methods for Canada and for 2020 and 2021.

| Year | Method | F<sub>1</sub>-score | Specificity | Sensitivity | Precision |
| --- | --- | --- | --- | --- | --- |
| 2020 | RF | 60.57 (59.97 - 61.18) | **98.80 (98.75 - 98.85)** | 48.50 (47.80 - 49.19) | **80.97 (80.29 - 81.66)** |
|  | XGB | **62.54 (61.99 - 63.09)** | 97.96 (97.89 - 98.03) | **54.41 (53.71 - 55.10)** | 73.77 (73.07 - 74.47) |
|  | LGB | 61.87 (61.36 - 62.39) | 98.17 (98.11 - 98.24) | 52.65 (52.03 - 53.27) | 75.24 (74.54 - 75.95) |
|  | RF SHAP | 61.72 (61.13 - 62.32) | 98.43 (98.37 - 98.50) | 51.39 (50.67 - 52.11) | 77.57 (76.83 - 78.31) |
|  | XGB SHAP | 61.65 (61.13 - 62.17) | 97.79 (97.72 - 97.86) | 54.03 (53.37 - 54.69) | 72.03 (71.27 - 72.79) |
|  | LGB SHAP | 61.48 (60.88 - 62.08) | 98.05 (97.97 - 98.12) | 52.72 (52.01 - 53.42) | 74.00 (73.20 - 74.80) |
| 2021 | RF | **63.81 (63.53 - 64.08)** | **98.76 (98.73 - 98.79)** | 52.03 (51.71 - 52.35) | **82.53 (82.15 - 82.92)** |
|  | XGB | 63.04 (62.75 - 63.32) | 98.18 (98.15 - 98.22) | **53.48 (53.16 - 53.79)** | 76.82 (76.43 - 77.21) |
|  | LGB | 61.33 (61.04 - 61.61) | 98.18 (98.14 - 98.22) | 51.41 (51.08 - 51.73) | 76.05 (75.65 - 76.46) |
|  | RF SHAP | 61.27 (60.97 - 61.57) | 98.57 (98.53 - 98.60) | 49.82 (49.48 - 50.15) | 79.64 (79.20 - 80.08) |
|  | XGB SHAP | 59.63 (59.33 - 59.93) | 98.11 (98.07 - 98.14) | 49.66 (49.32 - 50.01) | 74.68 (74.29 - 75.08) |
|  | LGB SHAP | 61.33 (61.03 - 61.62) | 98.18 (98.14 - 98.22) | 51.40 (51.07 - 51.73) | 76.07 (75.66 - 76.48) |

**Table SM3.** Performance metrics in % and the 95% CIs obtained by the proposed COVID-19 detection methods for Japan and for 2020 and 2021.

| Year | Method | F<sub>1</sub>-score | Specificity | Sensitivity | Precision |
| --- | --- | --- | --- | --- | --- |
| 2020 | RF | 56.85 (55.97 - 57.73) | **98.89 (98.82 - 98.97)** | 43.33 (42.45 - 44.21) | **83.22 (82.12 - 84.31)** |
|  | XGB | 59.25 (58.36 - 60.13) | 97.82 (97.72 - 97.92) | 49.51 (48.57 - 50.44) | 74.17 (73.04 - 75.30) |
|  | LGB | 71.54 (71.24 - 71.85) | 98.23 (98.19 - 98.26) | 63.99 (63.61 - 64.38) | 81.16 (80.81 - 81.50) |
|  | RF SHAP | 59.06 (58.28 - 59.84) | 97.95 (97.85 - 98.04) | 48.86 (48.04 - 49.68) | 75.03 (73.96 - 76.09) |
|  | XGB SHAP | 59.70 (58.83 - 60.58) | 97.82 (97.73 - 97.91) | 50.05 (49.13 - 50.98) | 74.31 (73.23 - 75.39) |
|  | LGB SHAP | **71.55 (71.25 - 71.85)** | 98.21 (98.17 - 98.24) | **64.09 (63.72 - 64.46)** | 81.03 (80.68 - 81.38) |
| 2021 | RF | 70.11 (69.85 - 70.38) | **98.95 (98.93 - 98.98)** | 59.24 (58.91 - 59.56) | **85.94 (85.61 - 86.27)** |
|  | XGB | 67.32 (67.03 - 67.60) | 98.48 (98.44 - 98.51) | 57.92 (57.58 - 58.25) | 80.42 (80.03 - 80.81) |
|  | LGB | 80.28 (80.22 - 80.34) | 78.53 (78.43 - 78.63) | **79.37 (79.29 - 79.44)** | 81.22 (81.14 - 81.30) |
|  | RF SHAP | 66.80 (66.54 - 67.06) | 98.55 (98.52 - 98.58) | 56.91 (56.58 - 57.25) | 80.92 (80.57 - 81.28) |
|  | XGB SHAP | 64.06 (63.77 - 64.34) | 98.40 (98.36 - 98.43) | 54.13 (53.80 - 54.47) | 78.50 (78.09 - 78.90) |
|  | LGB SHAP | **80.26 (80.21 - 80.32)** | 78.53 (78.42 - 78.64) | 79.34 (79.26 - 79.41) | 81.21 (81.13 - 81.30) |

**Table SM4.** Performance metrics in % and the 95% CIs obtained by the proposed COVID-19 detection methods for South Africa and 2020 and 2021.

| Year | Method | F<sub>1</sub>-score | Specificity | Sensitivity | Precision |
| --- | --- | --- | --- | --- | --- |
| 2020 | RF | 81.75 (81.54 - 81.95) | **92.55 (92.38 - 92.72)** | 78.06 (77.75 - 78.38) | 85.83 (85.54 - 86.12) |
|  | XGB | 80.19 (79.96 - 80.42) | 91.30 (91.13 - 91.48) | 77.03 (76.69 - 77.38) | 83.66 (83.36 - 83.95) |
|  | LGB | 79.66 (79.45 - 79.88) | 91.23 (91.06 - 91.40) | 76.27 (75.95 - 76.58) | 83.41 (83.13 - 83.69) |
|  | RF SHAP | **81.89 (81.68 - 82.10)** | 92.53 (92.36 - 92.70) | **78.31 (78.01 - 78.62)** | **85.84 (85.55 - 86.13)** |
|  | XGB SHAP | 77.42 (77.19 - 77.64) | 90.57 (90.36 - 90.77) | 73.49 (73.14 - 73.84) | 81.83 (81.50 - 82.17) |
|  | LGB SHAP | 79.55 (79.35 - 79.75) | 91.25 (91.08 - 91.42) | 76.07 (75.77 - 76.36) | 83.40 (83.12 - 83.68) |
| 2021 | RF | **77.70 (77.54 - 77.86)** | **91.83 (91.73 - 91.93)** | **72.47 (72.24 - 72.70)** | **83.76 (83.57 - 83.95)** |
|  | XGB | 74.87 (74.69 - 75.05) | 90.08 (89.96 - 90.20) | 70.06 (69.83 - 70.29) | 80.41 (80.18 - 80.64) |
|  | LGB | 73.83 (73.65 - 74.01) | 90.10 (89.98 - 90.22) | 68.50 (68.25 - 68.74) | 80.08 (79.86 - 80.31) |
|  | RF SHAP | 75.68 (75.52 - 75.84) | 90.70 (90.57 - 90.82) | 70.63 (70.42 - 70.84) | 81.53 (81.29 - 81.76) |
|  | XGB SHAP | 72.46 (72.28 - 72.64) | 89.52 (89.41 - 89.63) | 67.07 (66.82 - 67.32) | 78.81 (78.59 - 79.03) |
|  | LGB SHAP | 73.82 (73.64 - 74.00) | 90.14 (90.02 - 90.26) | 68.44 (68.19 - 68.69) | 80.13 (79.91 - 80.36) |

## Footnotes

1 Only for 2020

2 Only for 2021

